# Primary care transformation in Scotland: a comparison two cross-sectional national surveys of general practitioners’ views in 2018 and 2023

**DOI:** 10.1101/2024.08.14.24311940

**Authors:** E Donaghy, KD Sweeney, Ng Lauren, Holly Haines, Alexandra Thompson, D Henderson, HHX Wang, A Thompson, B Guthrie, SW Mercer

**Author notes:** Correspondence: Stewart Mercer, Usher Institute, University of Edinburgh, BioQuarter (Gate, 5-7, 3 Little France Rd, Edinburgh EH16 4UX.

## Abstract

**Objectives:** The new general practitioner (GP) contract for Scotland, introduced in 2018, established GP Clusters and expanded multidisciplinary team (MDT) working. This paper compares the views of GPs in Scotland regarding the new contract, their working lives, and career intentions in 2018 and 2023.

**Methods:** Cross-sectional postal survey of all qualified GPs in Scotland in late 2023 exploring views on Cluster working, MDT-expansion, their working lives, and career intentions, compared with a similar survey from 2018.

**Results:** Job pressure was significantly higher in 2023 than 2018, but overall job satisfaction and negative job attributes were unchanged, while positive job attributes improved. More GPs were planning to reduce their hours and leave direct patient care in 2023 than 2018.

Quality leads views on Cluster working were unchanged, with 70-80% reporting insufficient support in both surveys. Cluster knowledge and engagement was unchanged but all GPs showed small but significant increases in understanding of quality improvement. Most felt MDT expansion was insufficient to reduce workload and fewer GPs reported giving longer consultations for complex patients in 2023 than 2018. Significantly more practices were trying to recruit GPs in 2023, and GPs reported worsening NHS services, higher workload, and lower practice sustainability. Only 1 in 20 GPs in the 2023 survey thought that the new contract had improved the care of patients with multimorbidity.

**Conclusions:** GPs report few improvements in working life five years after the new contract was introduced, and are responding by planning to reduce their hours or leave direct patient care.

## Introduction

Internationally, ageing populations, rising multimorbidity, austerity, and widening inequalities are posing major challenges to health services.(1,2) General practice is fundamental to addressing these challenges.(3) Strong general practice is associated with lower health inequalities and healthcare costs(4,5) yet it is facing unprecedented crises globally.(6) There are concerns about the future of general practice in the UK, with shortages of general practitioners (GPs), declining continuity of care, and reduced access.(7) Internationally, responses to these challenges include reforms to manage increased demand whilst improving efficiency,(8) although frequently without appropriate investment.(9) A recent systematic scoping review found expansion of multidisciplinary teams (MDTs) was the most common reform in OECD countries.(10)

In 2014, the Scottish Government introduced legislation to integrate health and social care services, leading to the formation in 2016 of integrated authorities (IAs), and their delivery arm, Health and Social Care Partnerships (HSCPs).(11) In April 2018, the first-ever Scottish GP contract was introduced,(12) although elements of it began in 2016 when the Quality Outcomes Framework was abolished and GP clusters were introduced. Clusters are geographical groups of 5-8 practices working together to improve their local populations’ quality of care (intrinsic role) and provide local leadership within the IAs and HSCPs (extrinsic role).(12,13) Each practice has a Practice Quality Lead (PQL), and each cluster a Cluster Quality Lead (CQL). Clusters were expected to be functional by April 2017.(14,15) The 2018 contract aimed to reduce GP workload by expanding the MDT workforce, allowing GPs to focus on patients with complex needs as expert medical generalists.(12,13) As of March 2023, over 4700 whole-time equivalent new MDT staff were working in primary care in Scotland.(16)

A 2018 survey of GP clusters in Scotland reported a lack of support and training,(17) similarly echoed in subsequent qualitative interviews in 2020/2021 (18,19) Key barriers to effective cluster working included lack of time, poorly developed relationships, and limited data. Further interviews in 2022 with GPs and MDT staff found no perceived reduction in GP workload nor improvement in the care of patients with complex needs.(20) MDT staff reported challenges in building new relationships, adapting to patient complexity, and the fast pace of primary care. Issues over MDT line management, training and professional development needs were also highlighted.

A patient survey (n=1,053) in 12 Scottish practices across 3 Health Boards in 2022-2023 found patients in deprived-urban areas (compared with affluent-urban or remote and rural areas) had the most complex needs, but reported the poorest experience of GP consultations.(21) In-depth interviews with patients highlighted concerns about access, consultations length and continuity of care.(22) Further evaluation revealed limited patient awareness of MDT roles, and concerns about reception staff signposting to MDT-care, particularly for those with complex problems in high deprivation areas.(23)

## Methods

### Study design

Using the same process as the 2018 survey, (17,24) we conducted a postal survey of GPs in Scotland in 2023. In both years, the survey was posted to all qualified GPs.(25) In 2023, 4,529 surveys were sent in October 2023, with two reminders to non-responders. Data collection stopped in early March 2024. The response rate was 30%, compared to 56% in 2018,(24) although the characteristics of responding GPs were very similar in both years, and similar to national figures (supplementary table S1).

### Instruments used

The 2023 GP survey used the same validated measures of working life and future work intentions as the 2018 Scottish GP survey, as also used in the English GP working life biannual surveys since 1999 (26), with the most recent being in 2021.(27) (See supplementary file Box S1 for details of the questions asked). We additionally collected data on bespoke items on Cluster working, also collected in 2018,(17) and new items about MDT expansion based on our qualitative findings and a survey conducted by Public Health Scotland (20,28) as explained below.

#### Cluster variables

As described previously,(17) GP Quality Leads (QLs - both CQLs and PQLs) were asked about their experiences of Cluster meetings and level of support provided. All GPs were asked about their knowledge of and engagement with the Cluster and how it affected their knowledge of quality improvement (QI) (supplementary file Box S2).

#### MDT variables

The 2023 survey asked GPs which MDT staff they had access to in their practice, what impact these staff had on their workload, and what the advantages and disadvantages of MDT staff were. GPs were also asked a) what percentage of their previous clinical work was now delegated to MDT staff; b) what percent they felt could be safely delegated; and c) which staff would be most important if additional investment was available: more GPs, more MDT staff, or more administrative staff (supplementary file Box S3).

#### Additional questions

Information was collected on GP demographics and employment details. GPs were asked whether their practice scheduled longer GP appointments for patients with complex needs (e.g., those with multimorbidity and/or mental health problems) and if they felt the new GP contract was improving care for patients with multimorbidity who were either elderly or living in deprived areas. Questions from previous English GP Worklife surveys about job changes seen in the past 12 months were also asked in both years (supplementary file Box S4).

### Data analysis

The 2023 survey results are compared with the 2018 survey below. Given the similarity between the GP characteristics in both surveys (Supplementary Table S1), a direct statistical comparison was warranted. Standard parametric and non-parametric statistics were used, depending on the type and distribution of data for each variable. A Forest plot was drawn for the key variables which shows the effect sizes (Cohen’s d) of the differences between the 2023 and 2018 surveys.

## Results

Table 1 shows the working practices of the GPs who responded to the surveys. GPs reported working significantly fewer sessions per week and taking significantly fewer holidays per year in 2023 than in 2018.

**Table 1.** GP working practices in 2023 and 2018.

|  | **GP Survey 2023**<br>**(n=1378)** | **GP survey 2018**<br>**(n=2454)** | **P value** |
| --- | --- | --- | --- |
| Mean years in current practice (SD) | 12.2 (8.9) | 12.5 (9.4) | 0.951 |
| Mean sessions per week (SD) | 6.6 (1.6) | 6.9 (1.8) | <0.001 |
| Mean weeks holiday/year (SD) | 6.5 (1.6) | 6.8 (2.9) | <0.001 |
| Current employment model (n/%) |  |  |  |
| Partnership | 1143 (82.6%) | 2048 (83.8%) | 0.364 |
| Salaried | 232 (16.8%) | 387 (15.8%) |  |
| Locum/other | 8 (0.6%) | 10 (0.4%) |  |
| Future employment model (n/%) |  |  |  |
| Partnership | 957 (78.8%) | 1778 (79.6%) | 0.630 |
| Salaried | 203 (16.7%) | 351 (15.7%) |  |
| Locum/other | 55 (4.5%) | 106 (4.7%) |  |
| Actively seeking new model (n/%) | 87 (6.4%) | 160 (6.5%) | 0.873 |
| Does Out of Hours (n/%) | 222 (16.1%) | 585 (23.8%) | <0.001 |

### Current working life and future intentions

Table 2 shows the results for the domains of job satisfaction, job pressure, job attributes, and future work intentions. Although work pressures were significantly higher, mean positive job attributes significantly improved in 2023 and negative job attributes remained the same. Mean job satisfaction was similar in both survey years. For individual item scores for the four domains, see Supplementary Tables S2-S5.

**Table 2.** GPs current working life and future work intentions in 2023 and 2018.

|  | **GP Survey 2023**<br>**(n=1378)** | **GP survey 2018**<br>**(n=2454)** | **P value** |
| --- | --- | --- | --- |
| **Current working life:** |  |  |  |
| Job satisfaction | 5.31 (0.95) | 5.26 (0.99) | 0.128 |
| Job pressures | 3.52 (0.78) | 3.45 (0.74) | 0.004 |
| Positive job attributes | 3.34 (0.54) | 3.29 (0.59) | 0.011 |
| Negative job attributes | 3.85 (0.56) | 3.86 (0.58) | 0.594 |
| **Future intentions:** |  |  |  |
| *All GPs* |  |  |  |
| Increase hours | 1.41 (0.83) | 1.49 (0.95) | 0.005 |
| Decrease hours | 3.08 (1.56) | 2.81 (1.56) | <0.001 |
| Work outside UK | 1.44 (0.92) | 1.42 (0.85) | 0.354 |
| Leave direct patient care | 2.39 (1.53) | 2.24 (1.52) | 0.033 |
| Leave medical work entirely | 2.24 (1.52) | 2.18 (1.53) | 0.060 |
| *GPs aged <55 years* |  |  |  |
| Increase hours | 1.49 (0.88) | 1.59 (1.03) | 0.170 |
| Decrease hours | 2.74 (1.47) | 2.51 (1.44) | <0.001 |
| Work outside UK | 1.48 (0.94) | 1.44 (0.87) | 0.874 |
| Leave direct patient care | 1.91 (1.19) | 1.75 (1.11) | <0.001 |
| Leave medical work entirely | 1.72 (1.09) | 1.62 (1.04) | 0.002 |

Satisfaction items measured on a 7-point scale (1= extremely dissatisfied, 7= extremely satisfied). Work pressure is measured on a 5-point scale (1=no pressure, 5= high pressure). Job attributes are measured on a 5-point scale (1=strongly disagree, 5=strongly agree). Job control, job design, work pressure, and work load are derived from job attributes (see Methods). Future intentions measured on a 5-point scale: 1=none; 2=slight; 3=moderate; 4=considerable. 5=high

In 2023, significantly fewer GPs intended to increase their hours, and significantly more intended to decrease hours (44% in 2023 versus 36% in 2018) or leave direct patient care entirely (24% versus 22%) than in 2018 (Table 2). For GPs under 55 years of age, significantly more planned to reduce their hours (33% versus 26%), leave direct patient care (13% versus 10%), and leave medical work entirely (9% versus 8%) in 2023 than in 2018 (Table 2). Figure 1 shows the effect sizes of the mean scores for the measures shown in Table 2. Effect sizes were small, irrespective of their statistical significance.

**Figure 1.**
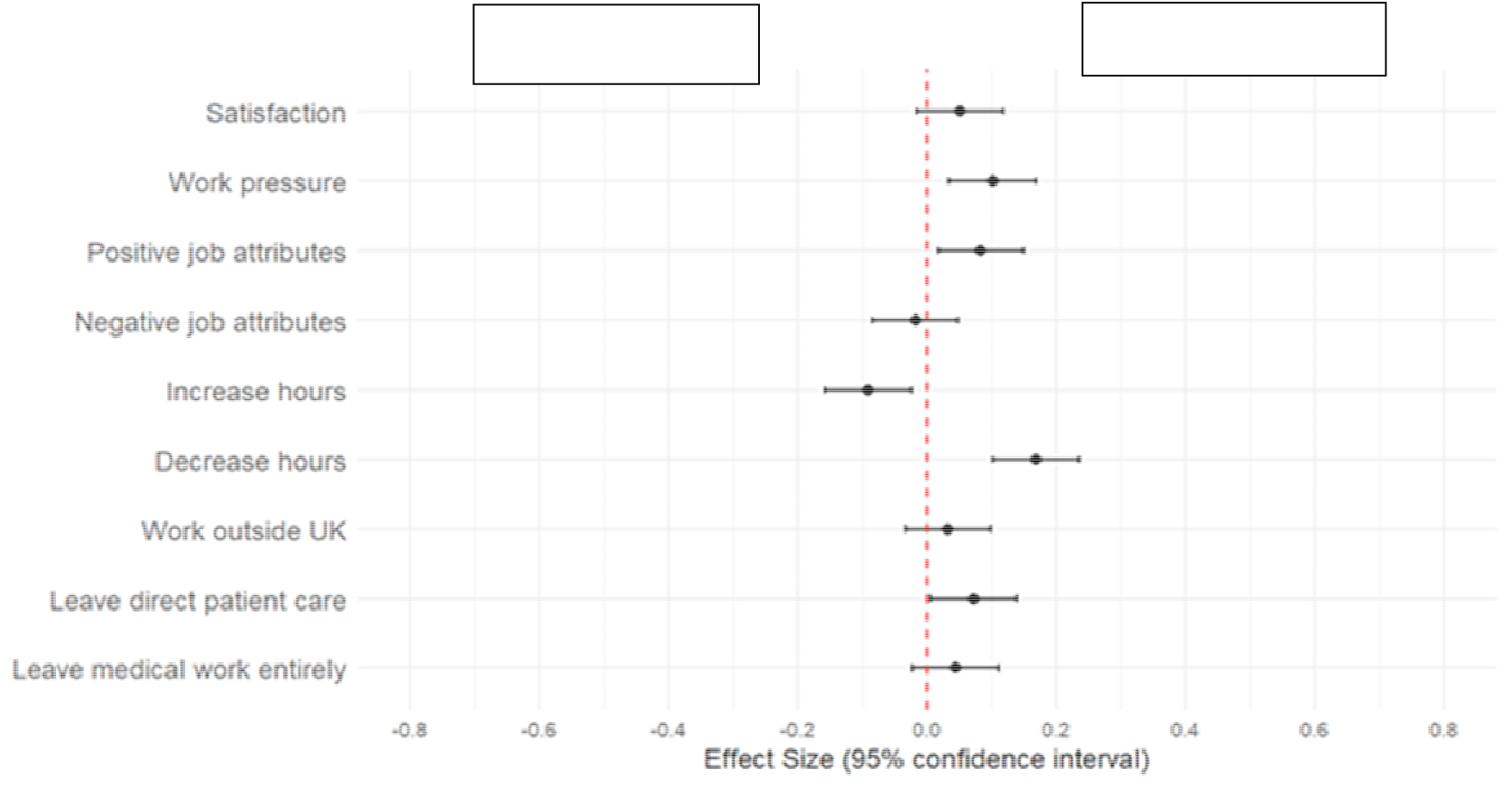
Effect size differences in working life domains and future work intentions of GPs in 2018 and 2023. Values that lie to the right of the red dotted vertical line mean that the scores were higher (better) in 2023 than in 2018, and those lying to the left mean the opposite. If the confidence intervals (black bars) cross the red line, then the difference was not statistically significant. It should be noted though that although statistically significant differences were found, the sizes of the differences were small (effect size of 0.2 or less).

### Clusters

QLs views on cluster meetings did not change between the two surveys (Figure 2a and Supplementary Table S6). There was no difference in the extent to which they felt supported overall (Figure 2a), but individual items showed significantly less support for analysis, and significantly more support for QI methods and leadership in 2023 than in 2018 (Supplementary Table S6). For all other GPs (neither CQLs nor PQLs), mean overall scores for knowledge and engagement with Clusters did not differ between 2023 and 2018, but significant improvements in individual items were found for ‘decisions’ and ‘queries’ (Figure 2b and Supplementary Table S6). There was a significant improvement in mean overall QI score in 2023 (p<0.001) which reflected increases in all six aspects of QI (Supplementary Table S6). Effect sizes were small for all measures (Supplementary Table S6).

**Figure 2.**
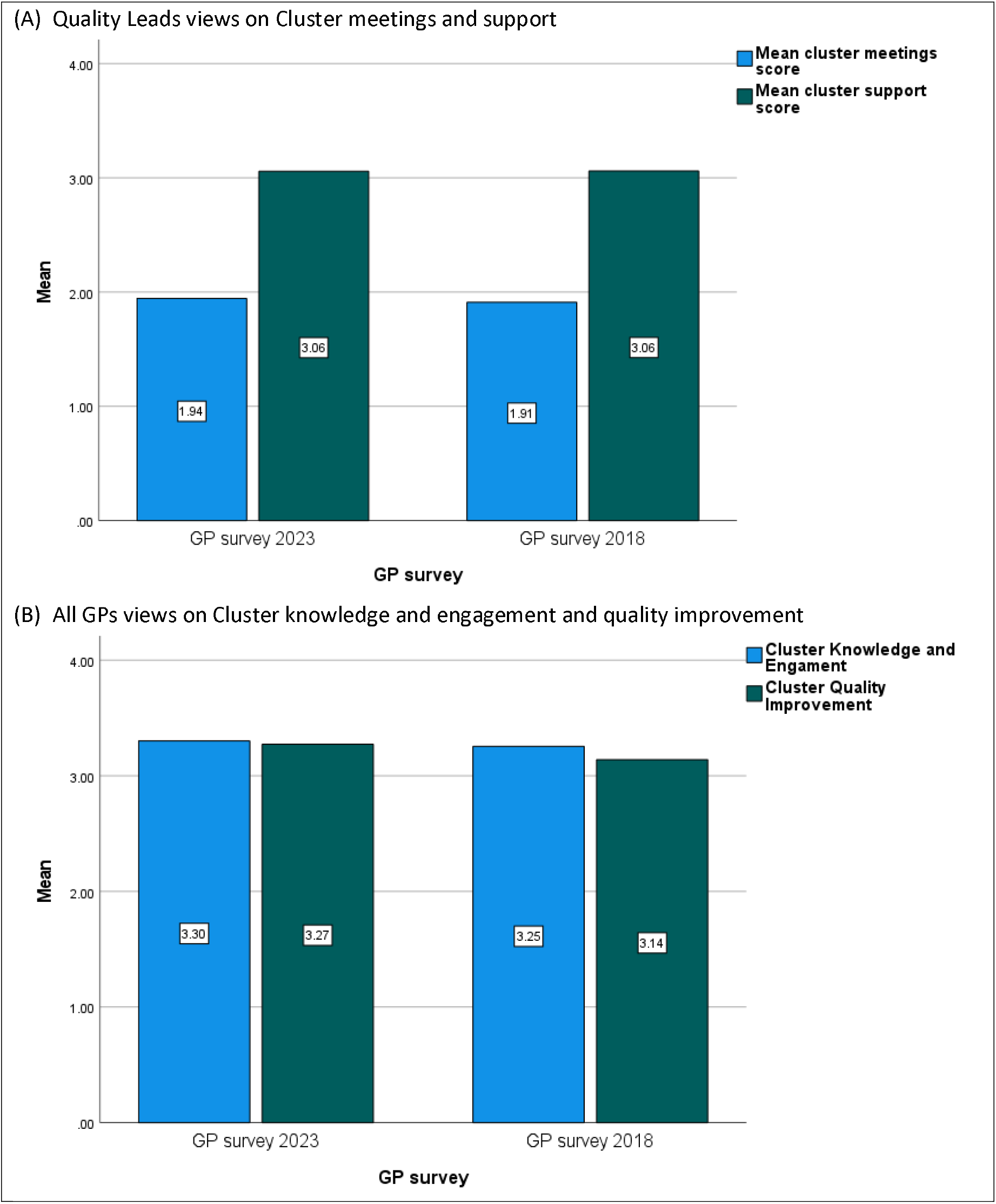
GPs views on Clusters in 2023 and 2018. Cluster meetings score measured on a 5-point scale; (1= always, 5 = never). Extent of support measured on a 4-point scale; 1=fully supported; 4=not at all supported. GPs knowledge about clusters scored on a 5-point scale; 1=strongly disagree, 5= strongly agree. Understanding of QI scored on a 5-point scale; 1=decreased a lot, 5= increased a lot (see Methods)

### Multidisciplinary teams

In 2023, GPs reported that 8.5% of workload had been delegated to MDT staff, but estimated that 22% could, in principal, be safely delegated (p<0.001). In 2023, 82% felt that more GPs was the most important issue for any future investment, compared with 69% in 2018 (p<0.001).

Table 3 shows the 2023 GPs’ views on new MDT staff (these questions were not asked in 2018). Access to different MDT staff varied widely. More than half of the GPs reported that access was insufficient to reduce their workload in all staff categories except vaccinations. 84% provided free-text answer about advantages of MDT expansion. 54.9% documented one positive comment, 28.9% two, and 9.8% three or more. One in 4 (23.4%) documented both positive and negative comments. For the positive comments, themes related to the additional clinical expertise and skill mix, better collaborative team working, learning from MDT staff, gaining new perspectives and ideas, better links with secondary care, better links with community resources, and improved patient care and patient safety. Comments on reduced workload was often qualified as ‘some reduction’ due to increased patient demand and insufficient MDT workforce. The negative comments largely related to the issues in the subsequent question about the disadvantages of the new MDT, including lack of clinical space (68% agreed), the need to provide training and supervision (60% agreed), and lack of control over what the MDT staff actually do (71% agreed) (Supplementary Table S7).

**Table 3.** GPs views on availability and impact on workload of new MDT staff - 2023.

|  | No access | Some access but insufficient to reduce my workload | Sufficient access to reduce my workload | Not sure |
| --- | --- | --- | --- | --- |
| Pharmacy | 16 (1.2%) | 778 (56.9%) | 579 (41.8%) | 1 (0.1%) |
| Urgent care | 570 (41.6%) | 406 (29.3%) | 390 (28.2%) | 10 (0.7%) |
| Advanced physiotherapy | 443 (32.1%) | 637 (46.1%) | 265 (19.1%) | 4 (0.3%) |
| Mental health | 477 (34.4%) | 637 (46.1%) | 265 (19.1%) | 5 (0.4%) |
| Link workers | 407 (29.5%) | 575 (41.6%) | 340 (24.6%) | 59 (4.3%) |
| CTAC nurses | 155 (11.2%) | 606 (43.9%) | 594 (43.0%) | 26 (1.9%) |
| Vaccinations | 153 (11.2%) | 223 (16.3%) | 834 (61.0%) | 157 (11.5%) |

CTAC: Community Treatment and Care Team

### Recruitment, local NHS services, sustainability, and training

More GPs in 2023 reported that their practices were trying to recruit GPs than in 2018 (35.8% versus 30.5%, p<0.01), with more trying for longer than 12 months (42% versus 30.9%, p<0.001). In 2023, more GPs felt that local NHS services had significantly worsened in the last 12 months, that practice and personal workload was higher, and that the long-term sustainability of their practice was worse, compared with 2018 (all p<0.001) (Supplementary Table S8).

Overall, more practices were involved in the training and education of undergraduate students in 2023 than 2018 (74% versus 59%, p<0.001). In 2023, 72% were involved in training (non-medical) healthcare professionals, compared with 55% in 2018 (p<0.001) (Supplementary file Figure S1).

### Improving the care of patients with complex needs

Fewer GPs reported giving longer consultations for complex patients in 2023 than in 2018 (39.8% versus 52.2%, respectively; p<0.001). Only 5% of GPs thought the new contract was improving the care of elderly patients with multimorbidity, and only 4% felt it was improving the care of multimorbid patients in deprived areas (Supplementary Figure S2).

## Discussion

### Summary of findings

Scottish GPs’ working lives, career intentions, and views on Cluster and MDT working in 2023 have changed relatively little since 2018. Job pressure is significantly higher but job satisfaction and negative job attributes are unchanged, while positive job attributes improved. However, more GPs are planning to reduce their hours and leave direct patient care than in 2018. Views on clusters were are largely unchanged but there are small improvements in understanding of QI. Less than half of the GPs reported that MDT expansion had significantly reduced their workload, and fewer reported offering longer consultations to complex patients than in 2018. Significantly more practices are trying to recruit GPs than in 2018, and GPs report worsening NHS services, higher workload, and lower practice sustainability. Only 1 in 20 GPs believed the new contract has improved care for patients with complex needs.

### Strengths and limitations

A key strength of this study is having comparable and broadly representative national data examining GP views across the first five years of the new Scottish GP contract. The 2023 response rate was lower than in 2018 (30% versus 56%), but the characteristics of responding GPs was virtually identical in both years. The limited impact of the new GP contract should be considered in the context of the disruption caused by the Covid-19 pandemic.

### Comparison with published literature

Our finding that job pressure has increased is mirrored by the 2021 English GP Worklife survey,(27) with other data from England showing a higher number of appointments being provided in 2023 than any year since 2018 despite a fall in the number of fully trained full-time equivalent GPs.(29,30) In contrast to our finding that overall job satisfaction in Scotland did not changed from 2018 to 2023, scores on the same measure in the English GP Worklife survey fell significantly between 2019 and 2021.(27) English data shows a continued trend of GPs reducing their working hours, and increasing numbers planning to leave direct patient care,(27,30) in keeping with our findings. We plan to compare the Scottish 2023 survey with next English GP Worklife survey (2023/4) when data becomes available.

Our findings on MDT staff echo those of a recent Public Health Scotland survey of GPs which reported insufficient access to MDT staff (particularly urgent care staff), and a lack of reduction in GP workload with no release of time for complex patients, and similar concerns about disadvantages.(31) Similar findings have been reported in England in the Additional Roles Reimbursement Scheme.(32)

### Implications for policy and practice

There is widespread recognition that primary care faces an unprecedented crisis, in the UK,) and in other high-income countries. This survey of Scottish GPs provides evidence for policy makers that recent reforms have not insulated general practice in Scotland from increasing workload and workforce pressures, despite this being an aim of the 2018 contract. This has implications for ongoing negotiations for phase two of the GP contract in Scotland, highlighting the need to take more robust measures to reduce GP workload and improve workforce sustainability.

## Conclusions

Although there have been small improvements in some aspects of GP working life in Scotland since the new contract in 2018, most aspects have remained the same and some have worsened, including work pressure. GPs - including younger GPs - are responding by planning to reduce their hours or leave direct patient care, which is a worrying picture given problems of GP recruitment and retention.

## Data Availability

All data produced in the present work are contained in the manuscript

## Acknowledgements

We would like to thank our Patient and Public Involvement group: Colin Angus (Chair), Morag Cullen, Mary Hemphill, Anne Marie Kennedy, who gave valuable feedback throughout the research programme. Special thanks to our Patient and Public Involvement Coordinator: Jayne Richards

We would like to thank all the GPs who contributed to the survey.

This study was funded through a research grant from the Economic and Social Research Council (reference: ES/T014164/1). Ethical approval was obtained from the Wales REC 6 research ethics committee (reference: 21/WA/0078), and research and development approval from participating Scottish Health Boards.

