## supplementary information and tables for "Primary care transformation in Scotland: a comparison two cross-sectional national surveys of general practitioners’ views in 2018 and 2023"

**Supplementary File**

**Box S1 – Details of working life questions**

***The working life survey*** has four related domains of working lives; job satisfaction, job stressors, positive job attributes, and negative job attributes, with each domain having a series of individual questions.[4] For each domain, we created an average score for all of the questions for each respondent.

Job satisfaction was measured by 10 different items with ratings on a 7-point scale from 1 “extremely dissatisfied” and to 7 “extremely satisfied”. The 10 answers were then averaged (minimum possible score of 1 and maximum possible score of 7).

Job stressors were measured as the pressure experienced from 15 job items, rated on a 5-point scale; 1 “no pressure”, 2 “slight pressure”, 3 “moderate pressure”, 4 “considerable pressure”, 5 “high pressure”. The 13 answers were averaged (minimum possible score of 1 and maximum possible score of 5).

Job attributes had eleven statements regarding ‘positive’ job attributes, and six statements regarding negative job attributes, where 1 indicates “strongly disagree”, 2 “disagree”, 3 “neutral”, 4 “agree” and 5 “strongly agree”. The answers for the positive and negative job attributes were separately averaged (minimum possible score of 1 and maximum possible score of 5).

GPs intentions (in the next 5 years) to reduce work hours, leave medical work entirely, leave direct patient care, or to continue medical work but outside the UK, were measured using a 5-point scale with 1 indicating “none”, 2 “slight”, 3 “moderate”, 4 “considerable” and 5 “high”.

**Box S2 – Details of Cluster questions**

As described previously [2], GP Quality Leads (QLs) – both Cluster Quality Leads (CQLs) and Practice Quality Leads (PQLs) were asked to what extent Cluster meetings were: 1) well organised; 2) friendly; 3) well facilitated; and 4) productive. They answered using a five-point response scale (‘always’, ‘nearly always’, ‘sometimes’, ‘hardly ever’, or ‘never’). QLs were also asked how supported they felt in relation to: 1) data; 2) health intelligence; 3) analysis; 4) quality improvement methods; 5) advice; 6) leadership; and 7) evaluation and research. They responded on a four-point scale (‘fully supported’, ‘almost fully supported’, ‘somewhat supported’, or ‘not at all supported’). They were also asked whether the focus was mainly on the ‘intrinsic’ role (quality improvement) or the ‘extrinsic role’ (participation in local planning of integrated care) of the Clusters.

All GPs were asked about their knowledge and engagement with the Cluster: 1) ‘I feel informed about what my Cluster is trying to achieve’; 2) ‘decisions made by my Cluster reflect my views’; 3) ‘when I make contact, my PQL is responsive to my queries and concerns’; 4) ‘my GP Cluster is “owned” by its members and feels like “our” organisation’; and 5) ‘I can influence the work of my cluster if I chose to’. The GPs rated their answers on a five-point scale from ‘strongly disagree;’ to ‘strongly agree’ (mid-point being ‘neutral’). All GPs were also asked how GP Clusters had affected their understanding of : 1) quality planning (how to set quality improvement goals); 2) quality improvement (methods and approaches); 3) quality control (measuring improvement, ensuring safety); 4) the characteristics of your local population of patients (such as age, deprivation, multimorbidity levels); 5) the quality of care they provide; and 6) the extent to which they involve patients in decisions about their care, based on what’s important to them. The answers were measured on a five-point scale from ‘decreased a lot’ to ‘increased a lot’ (mid-point ‘not changed’).

**Box S3 – Details of MDT questions**

GPs in the 2023 survey were asked about which MDT staff they had access to in their practice, and if this was sufficient to decrease their workload, which was rated as ‘no access’, ‘some access but insufficient to decrease my workload’; ‘sufficient access to reduce my workload’ and ‘don’t know’. They were then asked to describe the main advantages of the new MDT staff as free text. They were then asked if there had been any disadvantages of the new MDT staff in terms of clinical space, training and supervision, losing tasks they used to enjoy, and not having control over what the MDT actually do. Responses were on a seven-point Likert scale where 1 = ‘strongly agree’ and 7 = ‘strongly disagree’.

GPs were also asked a) what percentage of their previous clinical work was now delegated to MDT staff; b) what percent they felt could be safely delegated, and; c) which staff would be most important if additional investment was available – more GPs, more MDT staff, or more administrative staff.

**Box S4 – Additional Questions**

GPs were asked whether their practice scheduled longer GP appointments for patients with complex needs (such as those with multimorbidity and/or mental health problems) and if they felt the new GP contract was improving the care of patients with multimorbidity who were either; a) elderly or; b) living in deprived areas. These questions were all answered as ‘yes’; ’no’; or ‘don’t know’.

Information was also collected on GP demographics (age, gender, ethnicity), their model of employment, how long they had worked in the practice, the number of sessions they worked per week, holidays taken per year, and if they currently did out of hours work.

Questions from previous working life surveys in England were also asked (in both surveys) about changes they had seen in their job in the last 12 months in terms of NHS services locally, in their Practice, the practice workload, their personal workload, the sustainability of the practice, and their job satisfaction, on a 5 point Likert scale where 1 = ‘worsened a lot’ to 5 = ‘improved a lot’. They were also asked if their practice was trying to recruit more GPs (‘yes’; ‘no’; ‘don’t know’) and for how long (< 3 months; 3-6 months; 7-12 months; > 12 months). They were also asked their practice had been involved in training and education of a) undergraduate students; b) Foundation doctors; c) GPs in training; d) other healthcare professionals, and e) others.

Additional bespoke questions were included in the 2023 survey about social prescribing, green prescribing, and management of frail patients, which will be reported in a future paper.

**Table S1. Representativeness of GP survey and comparison with 2018 GP survey**

|  | **Scotland (2022)** | **GP survey (2023)** | **GP survey (2018)** |
| --- | --- | --- | --- |
| **Age (years)** |  |  |  |
| < 35 | 20% | 9% | 10% |
| 35-44 | 35% | 31% | 34% |
| 45-54 | 29% | 39% | 36% |
| 55-64 | 15% | 20% | 19% |
| 65 + | 1% | 1% | 1% |
| **Gender** | 61% | 57% | 58% |
| **Ethnicity (white)** | - | 92% | 91% |
| **GP Partner** | 72% | 83% | 84% |
| **Deprivation** | 20% | 20% | 20% |
| **Urban locality** | 73% | 69% | 65% |

Deprivation is calculated as the proportion of patients in the most deprived quintile of all practices in Scotland, calculated from the percentage of registered patients who are living in the top 15% most deprived areas based on the Scottish Index of Multiple Deprivation (SIMD). Urban locality is based on the proportion of registered patients living in urban conurbations as defined by the Scottish Government.

**Table S2. Individual domains of GP job satisfaction in 2023 and 2018**

|  | **GP survey 2023** | **GP survey 2018** | P value | Effect size |
| --- | --- | --- | --- | --- |
| **Satisfaction with:** Mean (SD) |  |  |  |  |
| Physical working conditions | 5.42 (1.42) | 5.49 (1.46) | 0.018 | -0.047 |
| Freedom to choose method of working | 5.48 (1.21) | 5.33 (1.29) | 0.002 | 0.118 |
| Colleagues and fellow workers | 6.18 (1.18) | 6.09 (1.15) | 0.016 | 0.081 |
| Recognition for good work | 4.66 (1.56) | 4.72 (1.55) | 0.167 | -0.043 |
| Amount of responsibility | 5.45 (1.47) | 5.48 (1.45) | 0.460 | -0.022 |
| Remuneration | 4.77 (1.63) | 4.92 (1.58) | 0.004 | -0.098 |
| Opportunities to use abilities | 5.47 (1.27) | 5.36 (1.27) | 0.004 | 0.081 |
| Hours of work | 4.75 (1.71) | 4.51 (1.68) | <0.001 | 0.140 |
| Variety in job | 5.74 (1.17) | 5.59 (1.23) | 0.001 | 0.128 |
| Overall feelings about job | 5.07 (1.24) | 5.01 (1.35) | 0.630 | 0.049 |

Satisfaction items measured on a 7-point scale (1= extremely dissatisfied, 7= extremely satisfied).

**Table S3. Individual domains of GP job pressure in 2023 and 2018**

|  | **GP Survey 2023** | **GP survey 2018** | P value | Effects size |
| --- | --- | --- | --- | --- |
| **Pressure from:** Mean (SD) |  |  |  |  |
| Increased patient demand | 4.10 (0.89) | 4.0 (0.92) | 0.001 | 0.110 |
| Problem patients | 3.82 (0.95) | 3.78 0.97) | 0.433 | 0.037 |
| Early discharge from hospital | 3.71 (1.04) | 3.58 (0.99) | <0.001 | 0.129 |
| Patient complaints | 3.28 (1.17) | 3.19 (1.14) | 0.023 | 0.078 |
| Insufficient time | 3.93 (1.04) | 3.91 (1.14) | 0.418 | 0.016 |
| Interruptions during consultations | 2.80 (1.14) | 2.62 (1.10) | <0.001 | 0.161 |
| Unrealistic expectations | 3.49 (1.22) | 3.30 (1.22) | <0.001 | 0.157 |
| Insufficient resources | 3.43 (1.21) | 3.19 (1.17) | <0.001 | 0.203 |
| Long hours | 3.49 (1.21) | 3.48 (1.16) | 0.836 | 0.000 |
| Paper work | 3.77 (1.04) | 3.78 (0.99) | 0.852 | -0.004 |
| External bodies | 3.01 (1.10) | 3.03 (1.05) | 0.366 | -0.025 |
| Quality linked payments | 2.47 (1.11) | 2.59 (1.06) | <0.001 | -0.112 |
| Finding locums | 3.44 (1.42) | 3.61 (1.43) | <0.001 | -0.114 |
| Negative media | 3.86 (1.17) | 3.33 (1.25) | <0.001 | 0.435 |
| Increasing workload | 4.24 (0.93) | 4.16 (0.94) | <0.001 | -0.093 |

Work pressure is measured on a 5-point scale (1=no pressure, 2=slight pressure, 3=moderate pressure, 4=considerable, 5= high pressure)

**Table S4. Individual domains of GP positive job attributes in 2023 and 2018**

|  | **GP survey2023** | **GP survey 2018** | **P value** | **Effect size** |
| --- | --- | --- | --- | --- |
| **Positive job attributes: M**ean (SD) |  |  |  |  |
| Choice in how I do my job | 3.63 (0.87) | 3.47 (1.00) | <0.001 | 0.167 |
| Choice in what I do at work | 3.20 (0.97) | 3.14 (1.06) | 0.014 | 0.067 |
| Working time is flexible | 2.88 (0.97) | 2.87 (1.19) | 0.340 | 0.004 |
| Interesting variety in things | 3.99 (0.75) | 4.03 (0.81) | 0.600 | -0.045 |
| Involved in deciding on changes | 3.36 (1.14) | 3.19 (1.17) | <0.001 | 0.151 |
| I always know my work responsibilities | 3.92 (0.81) | 3.79 (0.91) | <0.001 | 0.148 |
| Consulted about changes | 3.19 (1.17) | 3.09 (1.19) | <0.001 | 0.085 |
| Clear feedback about performance | 2.78 (0.98) | 2.74 (1.03) | 0.027 | 0.044 |
| I decide how to do my work | 3.21 (0.10) | 3.19 (1.07) | 0.074 | 0.025 |
| Changes have led to better patient care (D) | 2.52 (1.04) | 2.62 (1.05) | -0.098 | 0.004 |
| Patients trust my generalist skills | 4.01 (0.64) | 4.07 (0.74) | -0.084 | 0.014 |

Job attributes are measured on a 5-point scale (1=strongly disagree, 2=disagree, 3=neutral, 4=agree, 5=strongly agree)

**Table S5. Individual domains of GP negative job attributes in 2023 and 2018**

|  | **GP survey 2023** | **GP survey 2018** | **P value** | **Effect size** |
| --- | --- | --- | --- | --- |
| **negative job attributes:** Mean (SD) |  |  |  |  |
| I have to work very fast | 4.14 (0.82) | 4.11 (0.89) | 0.092 | -0.041 |
| I have to work very intensively | 4.34 (0.755) | 4.37 (0.792) | 0.787 | 0.034 |
| Patient complexity has increased | 4.71 (0.52) | 4.68 (0.42) | 0.028 | -0.054 |
| I do not have time to do all my work | 3.79 (1.09) | 3.81 (1.09) | 0.750 | 0.019 |
| Relationships at work are strained | 2.25 (1.05) | 2.28 (1.10) | 0.502 | 0.028 |
| I am required to do unimportant tasks | 3.88 (1.00) | 3.93 (1.02) | 0.330 | 0.055 |

Job attributes are measured on a 5-point scale (1=strongly disagree, 2=disagree, 3=neutral, 4=agree, 5=strongly agree)

**Table S6. GPs views on Clusters in 2023 and 2018**

| Mean (SD) | **GP survey 2023** | **GP survey 2018** | **P value** | **Effect size** |
| --- | --- | --- | --- | --- |
| **Quality Leads views** |  | | | |
| *Cluster meetings are:* |  | | | |
| Well organised | 1.94 (0.82) | 1.96 (0.84) | 0.735 | 0.028 |
| Friendly | 1.43 (0.62) | 1.47 (0.59) | 0.233 | 0.072 |
| Productive | 2.53 (1.00) | 2.50 (0.96) | 0.648 | - 0.028 |
| Well-facilitated | 1.97 (0.90) | 2.01 (0.87) | 0.376 | 0.042 |
| Total average score for meetings | 1.95 (0.68) | 1.98 (0.61) | 0.578 | -0.039 |
| *Cluster activities:* |  | | | |
| Focus om intrinsic role | 2.36 (0.88) | 2.24 (0.80) | 0.083 | -0.143 |
| Focus on extrinsic role | 2.97 (1.58) | 2.75 (1.01) | 0.062 | -0.173 |
| *Extent of support in relation to*: |  |  |  |  |
| Data | 3.02 (0.80) | 2.94 (0.753) | 0.051 | -0.110 |
| Health intelligence | 3.15 (0.82) | 3.09 (0.73) | 0.068 | -0.083 |
| Analysis | 3.22 (0.81) | 3.12 (0.72) | 0.006 | -0.138 |
| QI methods | 2.92 (0.80) | 3.05 (0.74) | 0.041 | 0.160 |
| Advice | 2.92 (0.82) | 3.00 (0.74) | 0.191 | 0.109 |
| Leadership | 2.85 (0.85) | 3.00 (0.81) | 0.009 | 0.183 |
| Evaluation and research | 3.29 (0.72) | 3.29 (0.69) | 0.831 | 0.001 |
| Total average score for support | 3.07 (0.66) | 3.06 (0.63) | 0.578 | -0.007 |
| **All other GPs views** |  |  |  |  |
| *Cluster knowledge and engagement:* |  |  |  |  |
| Informed about Cluster | 3.27 (1.11) | 3.25 (1.11) | 0.488 | 0.019 |
| Decisions reflect my views | 3.19 (0.93) | 3.10 (0.95) | 0.003 | 0.093 |
| PQL responsive to queries | 3.74 (0.87) | 3.64 (0.86) | <0.001 | 0.115 |
| Cluster owned by its members | 3.09 (1.00) | 3.06 (1.00) | 0.328 | 0.031 |
| Can influence work of Cluster | 3.23 (1.03) | 3.23 (1.00) | 0.876 | 0.003 |
| Total average score CKE | 3.30 (0.82) | 3.25 (0.89) | 0.082 | 0.059 |
| *Understanding of QI:* |  |  |  |  |
| Quality planning | 3.30 (0.67) | 3.14 (0.69) | <0.001 | 0.232 |
| Quality improvement | 3.32 (0.67) | 3.15 (0.68) | <0.001 | 0.249 |
| Quality control | 3.29 (0.64) | 3.12 (0.65) | <0.001 | 0.265 |
| Local population | 3.32 (0.66) | 3.21 (0.65) | <0.001 | 0.162 |
| Quality of care | 3.27 (0.60) | 3.13 (0.55) | <0.001 | 0.245 |
| Shared-decision making with patients | 3.16 (0.43) | 3.23 (1.00) | <0.001 | 0.150 |
| Total average score CQI | 3.27 (0.82) | 3.14 (0.51) | <0.001 | 0.260 |

Cluster meetings and activities items are measured on a 5-point scale; (1= always, 2= nearly always, 3=sometimes, 4=hardly ever, 5 = never). Extent of support measured on a 4-point scale; 1=fully supported; 2=almost fully supported; 3=somewhat supported; 4=not at all supported. Other GPs knowledge about clusters scored on a 5 point scale; 1=strongly disagree, 2=disagree, 3=neutral, 4=agree, 5= strongly agree. Understanding of QI scored on a 5 point scale; 1=decreased a lot, 2=decreased a little, 3=not changed, 4= increased a little, 5= increased a lot

**Table S7. Disadvantages of new MDT staff**

|  | Strongly agree | Agree | Slightly agree | Neutral | Slightly disagree | Disagree | Strongly disagree |
| --- | --- | --- | --- | --- | --- | --- | --- |
| Lack of clinical space | 316 (22.9%) | 303 (22.0%) | 319 (23.2%) | 133 (9.7%) | 51  (3.7%) | 172 (12.5%) | 83  (6.0%) |
| Training and supervision | 200 (14.6%) | 330 (24.1%) | 308 (22.4%) | 200 (14.6%) | 78  (5.7%) | 184 (13.4%) | 72  (5.2%) |
| Taken over tasks I enjoyed | 78  (5.7%) | 195 (14.2%) | 181 (13.2%) | 249 (18.1%) | 165 (12.0%) | 341 (24.8%) | 165 (12.0%) |
| Lack of control over their job | 483 (35.2%) | 301 (21.9%) | 194 (14.1%) | 187 (13.6%) | 62  (4.5%) | 88  (6.4%) | 59  (4.3%) |

**Table S8. GPs views in changes in the last 12 months in 2023 and 2018**

| Mean (SD) | **GP survey 2023** | **GP survey 2018** | **P value** | **Effect size** |
| --- | --- | --- | --- | --- |
| NHS services for your patients | 2.04 (1.09) | 2.37 (1.02) | **<0.001** | 0.319 |
| NHS services in your local area | 1.60 (0.84) | 2.00 (0.89) | **<0.001** | 0.448 |
| Practice workload | 1.78 (0.82) | 2.03 (0.87) | **<0.001** | 0.296 |
| Personal workload | 1.93 (0.91_ | 2.11 (0.93) | **<0.001** | 0.195 |
| Practice sustainability | 2.26 (1.03) | 2.45 (1.03) | **<0.001** | 0.185 |
| Job satisfaction | 2.43 (1.00) | 2.58 (0.93) | **<0.001** | 0.150 |

Measured on a 5-point scale; 1= worsened a lot, 2= worsened a little, 3= not changed, 4=improved a little, 5= improved a lot.

**Figure S1. GP practice involvement in education and training in 2023 and 2018**

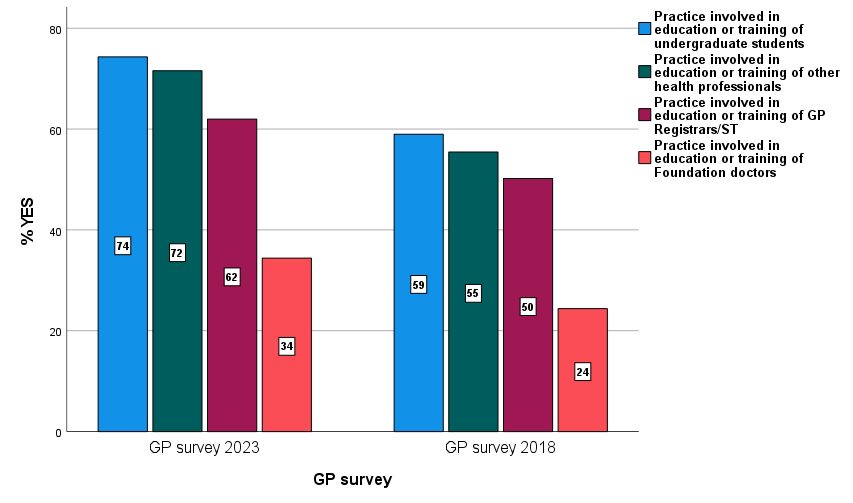

P<0.001
